# Risk Factors for Large Cell Transformation in Patients with Sezary Syndrome

**DOI:** 10.1101/2020.06.01.20119669

**Authors:** Neil K. Jairath, Ruple Jairath, Redina Bardhi, John S. Runge, Ramona Bledea, Yang Wang, Alexandra Hristov, Ryan A. Wilcox, Lam C. Tsoi, Matthew Patrick, Trilokraj Tejasvi

## Abstract

**Objectives:** Large cell transformation (LCT) of Sezary Syndrome (SS) is associated with an aggressive clinical course. To date, there are no rigorous studies identifying risk factors for the development of this phenomenon. We aim to characterize the clinicopathologic risk factors that may predispose patients with SS to develop LCT in the largest such study to date that the authors have identified.

**Materials/Methods:** We retrospectively evaluated all SS patient records available in the Michigan Medicine Cancer Registry from 2010–2019. The Mann-Whitney U test and Fisher exact test were used to compare age, sex, race, time to diagnosis, stage, total body surface area (TBSA) involvement, pathologic features, complete blood counts, flow cytometry data, and T cell receptor rearrangements. The Kaplan-Meier method and log-rank test were used to assess overall survival (OS). Univariate analyses were conducted for endpoints of LCT and OS and visualized with Forest plots using the “survival” and “forestplot” packages in R.

**Results:** Of the 28 SS patients included in the analysis, eight patients with LCT were identified, and 20 without nonlarge cell transformation (NLCT). Mean peak LDH before LCT (p = 0.0012), mean maximum TBSA involvement before diagnosis of LCT (p = 0.0114), absolute CD8^+^ cell count on flow cytometry or on biopsy at diagnosis of SS (p = 0.0455), presence of Langerhans cell hyperplasia (p = 0.0171), and presence of ulceration on biopsy (p = 0.0034) were clinicopathologic variables identified as differing significantly between the two groups. On univariate analysis, increased TBSA involvement (HR 1.043 per unit increase, 95% CI 1.001 – 1.081, p = 0.018) and increased peak LDH prior to LCT diagnosis (HR 1.002 per unit increase, 95% CI 1.001 – 1.003, p = 0.002) were identified as poorly prognostic, while unit increase in CD8^+^ absolute cell count at diagnosis of SS (HR 0.988, 95% CI 0.976 – 0.999, p = 0.041) was identified as protective for development of LCT. There was no survival difference identified between patients with “High” vs. “Low” CD8^+^ cell counts, or between LCT and NLCT groups.

**Conclusions:** Maximum TBSA involvement, peak LDH, presence of ulceration, Langerhans cell hyperplasia, and decreased levels of CD8^+^cells in the peripheral blood may predict the development of LCT in patients with SS.

## Introduction

Séary syndrome constitutes less than 5% of all cutaneous T cell lymphomas (CTCL) cases [1]. It is understood to be the “leukemic variant of mycosis fungoides (MF)” [1]. According to the WHO/EORTC classification of cutaneous lymphomas, it is characterized by erythroderma, generalized lymphadenopathy, and the presence of neoplastic T‐cells (Sézary cells) in skin, lymph nodes, and peripheral blood [2]. Sezary syndrome is associated with poor survival and progresses rapidly [3, 4], with a 5-year survival rate of approximately 11% [3, 4]. In contrast, MF represents approximately 50% of all primary cutaneous lymphomas, and thus is the most prevalent primary CTCL [2, 5]. Small to medium-sized T cells with cerebriform nuclei compromise MF lesions [2]. As opposed to the aggressive nature of SS, MF progresses slowly from thin patches to tumors over many years, even decades [2].

Patients with MF or SS may undergo transformation and develop large cell lymphoma [5]. This has been termed “large cell transformation” (LCT) and is defined by “the presence of large cells (CD30 +/−) exceeding 25% of the infiltrate throughout the lesion or forming microscopic nodules of large cells” [5, 6]. It occurs in 20% of advanced CTCL cases and is associated with aggressive behavior [7]. The median survival time for patients with LCT is 19–36 months [8, 9]. At present, the clinicopathologic features and underlying molecular mechanisms involved in the progression to LCT are not well understood [7].

Given the paucity of information in the reported literature concerning the LCT risk factors for patients with SS due to its low incidence, and the generally poor prognosis associated with this transformation, there is a need to reveal the associated factors and to assess the risk of LCT before it is diagnosed. This knowledge is essential to developing targeted therapies for these patients, and preventing the complications associated with more aggressive disease courses. Here, we seek to retrospectively characterize the clinicopathologic risk factors that may predispose patients with SS to LCT.

## Methods

This study was conducted with the approval of the Institutional Review Board at the University of Michigan. We retrospectively evaluated all SS patient records available at the Michigan Medicine from 2010–2019. The database of patients was queried using the following Boolean search parameters: free text including ‘large cell’, or ‘progression’ with a diagnosis of ‘Sezary Syndrome’, ‘mycosis fungoides’ or ‘cutaneous T-cell lymphoma’. Records were independently reviewed by N.K.J. and T.T. to confirm diagnosis of SS in all included patients and LCT in the appropriate patient subset. Patients must have had at least one histopathologically confirmed biopsy for each of the indicated diagnoses to be included in their respective study groups, reviewed by a dermatopathologist with expertise in cutaneous lymphoma to confirm the findings (A.H.). Cases which met the above criteria were then clinically confirmed as SS by N.K.J., R.B., and T.T. via detailed record review and integration of immunohistochemical and molecular data in consensus with clinical cutaneous lymphoma experts in dermatology and hematology (T.T. and R.A.W.).

All patients were classified according to the International Society for Cutaneous Lymphomas (ISCL) and European Organization of Research and Treatment of Cancer (EORTC) revised criteria of 2007 [10]. Staging included physical examination, peripheral blood smear, flow cytometry, blood cell count and chemistry, and computed tomography (CT) or positron emission tomography (PET) scan of the chest, abdomen, and pelvis.

### Clinical and pathological data

Patient charts, imaging, histopathology slides, molecular data, and pathology reports were all reviewed. The accompanying prognostic factors were collected and analyzed (full details in **Supplemental Table 1**): age at diagnosis at each of CTCL, SS, and LCT, sex, race/ethnicity, clinical stage at first diagnosis, date of each diagnosis, clinical findings at first diagnosis, first histological documentation of CTCL, SS, and LCT, dates of death or last follow up, location and number of skin sites biopsied, maximum total body surface area (TBSA) involvement before LCT, serum lactate dehydrogenase prior to and at LCT, complete blood count at diagnosis of Sezary Syndrome, diagnosis of LCT, and most recently, evidence of clonal T-cell rearrangements in tissue and/or blood samples, medication and treatment history specific to CTCL and SS, as well as treatment duration and response. Response to treatment was classified as either complete remission, partial response, stable disease, or progressive disease, assessed clinically according to the ISCL/EORTC guidelines [10].

The following pathological features were reviewed and documented (Full details in **Supplemental Table 1**): dates and locations of all recorded and reviewed biopsies, presence of LCT, presence of abnormal T-cells, presence/absence of folliculotropism, fibrosis, ulceration, epidermotropism, Pautrier microabscesses, spongiosis, neutrophil infiltration, Langerhans cell hyperplasia, and follicular mucin, flow cytometry data at diagnosis of Sezary Syndrome, diagnosis of LCT, and most recently, including absolute and relative values for CD1a, CD2, CD3, CD4, CD4+CD7-, CD4+CD26-, CD4+CD7-CD26-, CD5, CD7, CD8, CD10, CD16,56, CD19, CD20, CD45, CD56, NK Cells, and the CD4:CD8 ratio.

### Statistical Analysis

Analyses were carried out using software packages from R Foundation for Statistical Computing (Vienna, Austria). The Mann-Whitney U test, Fisher’s Exact Test, and Mantel-Cox analysis were used to compare variables. The Kaplan-Meier method was used to assess overall survival (OS) and differences were compared using the Mantel-Cox long rank test. Univariate analyses were conducted and visualized with Forest plots using the “survival” and “forestplot” packages in R. Overall survival (OS) was defined as the time from CTCL diagnosis to death, unless otherwise stated. For continuous variables analyzed using the Kaplan-Meier method, namely absolute CD8^+^ T cell count, the median value for all patients in the dataset was calculated, and designations of “High” or “Low” were given based on whether the value for each individual patients was higher or lower than the sample median, respectively. *p* values of <0.05 were considered statistically significant.

## Results

### Clinical Features

Medical records from 794 entries from 337 unique patients were retrieved in the initial collection using the aforementioned search strategy. Of these, only 85 patients were found to have either SS or LCT diagnosis. The authors manually reviewed these 85 patient records and excluded any patients that did not have a retrievable confirmed histopathologic diagnosis of SS. After applying exclusion criteria, 28 patients were included in the final analysis (**Figure 1**). The median age at diagnosis of SS was 72 (range 46 – 99). The diagnosis of SS preceded or was concurrent with the diagnosis of LCT in all cases in which transformation occurred. At the time of data analysis, 18 of the 28 (64%) patients in the study had died, and 10 (36%) were alive. One patient had a history of metastatic lung adenocarcinoma, concurrent with their CTCL diagnosis, and one patient had a history of metastatic melanoma, also concurrent with their CTCL diagnosis. Concise demographic tables and summary statistics can be visualized in **Table 1A**. Full demographic details can be visualized in **Supplemental Table 1**.

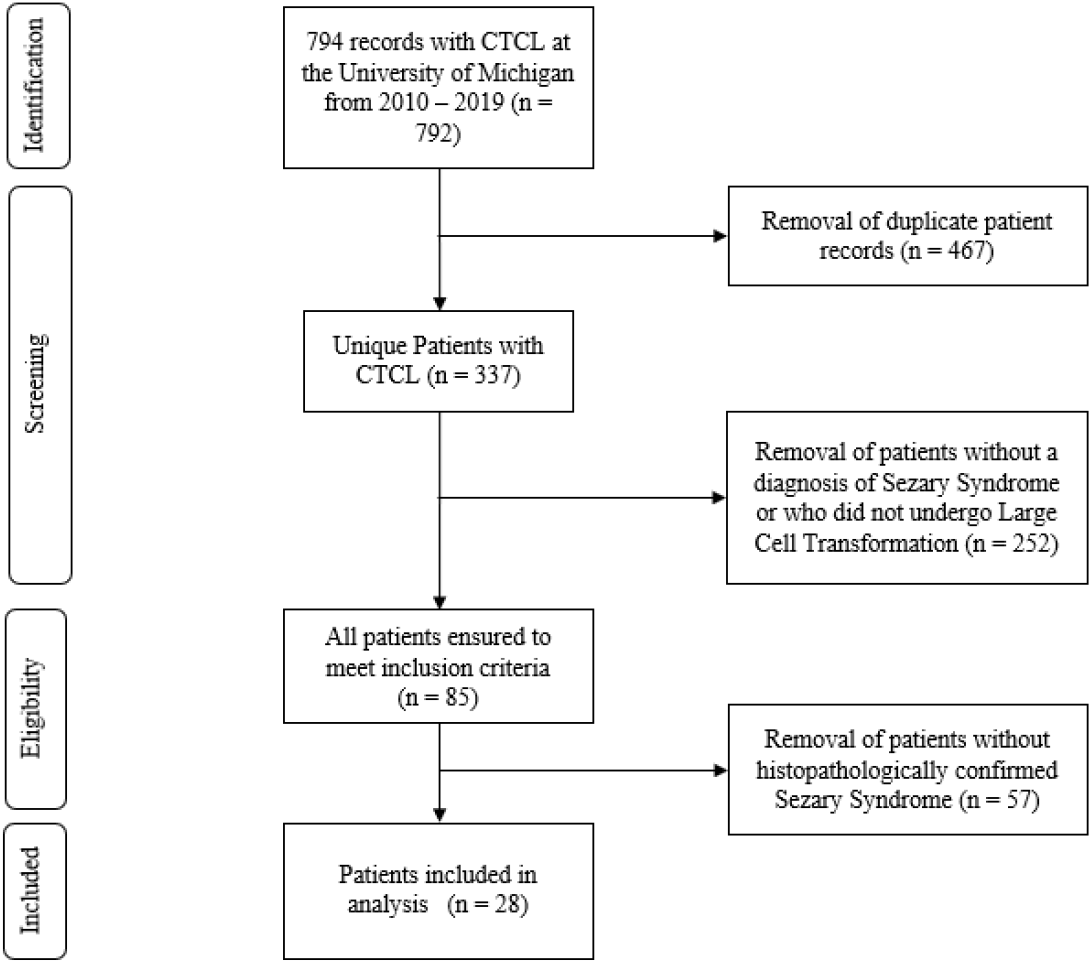
Figure 1:

**Table 1A:** Concise demographic details for each patient included in the study

| NLCT | Status | Days from CTCL Dx to Death | Days from SS Dx to death | Sex | Race | Stage at dx | Age at CTCL dx |
| --- | --- | --- | --- | --- | --- | --- | --- |
| Patient 1 | Dead | 2692 | 69 | Female | Caucasian | 1B | 63 |
| Patient 2 | Dead | 847 | 847 | Female | Caucasian | 1B | 72 |
| Patient 3 | Dead | 459 | 396 | Male | Caucasian | 3A | 72 |
| Patient 4 | Dead | 2275 | 2275 | Male | Caucasian | 3B | 84 |
| Patient 5 | Dead | 1561 | 1558 | Female | Caucasian | 3B | 88 |
| Patient 6 | Dead | 158 | 158 | Female | African American | 4 | 91 |
| Patient 7 | Alive | NA | NA | Male | Caucasian | 3B | 58 |
| Patient 8 | Alive | NA | NA | Male | Caucasian | 1B | 70 |
| Patient 9 | Alive | NA | NA | Female | Caucasian | 4A | 73 |
| Patient 10 | Alive | NA | NA | Female | Caucasian | 3 | 74 |
| Patient 11 | Alive | NA | NA | Male | Caucasian | 3 | 76 |
| Patient 12 | Dead | 338 | 338 | Female | Caucasian | 3 | 76 |
| Patient 13 | Dead | 1190 | 997 | Female | Caucasian | 3A | 81 |
| Patient 14 | Dead | 247 | 239 | Male | Caucasian | 3A | 74 |
| Patient 15 | Alive | NA | NA | Male | Caucasian | 3B | 65 |
| Patient 16 | Alive | NA | NA | Female | African American | 3A | 99 |
| Patient 17 | Alive | NA | NA | Male | Caucasian | 1B | 60 |
| Patient 18 | Dead | 353 | 326 | Female | Caucasian | 3A | 56 |
| Patient 19 | Dead | 336 | 127 | Male | Caucasian | 1B | 74 |
| Patient 20 | Alive | NA | NA | Female | Caucasian | 3A | 71 |
| LCT | Status | Days from CTCL Dx to Death | Days from SS Dx to death | Sex | Race | Stage at dx | Age at CTCL dx |
| Patient 21 | Dead | 663 | 419 | Male | Caucasian | 1B | 71 |
| Patient 22 | Alive | NA | NA | Male | Caucasian | 3 | 74 |
| Patient 23 | Dead | 952 | 952 | Male | Caucasian | 3 | 48 |
| Patient 24 | Dead | 1803 | 1365 | Female | Caucasian | 3B | 88 |
| Patient 25 | Dead | 6548 | 550 | Male | Caucasian | 2B | 59 |
| Patient 26 | Dead | 1631 | 1631 | Male | Caucasian | 3 | 74 |
| Patient 27 | Dead | 788 | 736 | Female | African American | 2B | 46 |
| Patient 28 | Dead | 444 | 444 | Female | Caucasian | 4A | 47 |
| Summary Statistics | Status | Days from CTCL Dx to Death | Days from SS Dx to death | Sex | Race | Stage at dx | Age at CTCL dx |
| NLCT Number | 11/9* |  |  | 9/11* | 18/2* | 5/13/2* |  |
| LCT Number | 7/1* |  |  | 5/3* | 7/1* | 3/4/1* |  |
| NLCT Average |  | 950.5 | 666.4 |  |  |  | 74 |
| LCT Average |  | 1832.7 | 871 |  |  |  | 63 |
| NLCT Max |  | 2692 | 2275 |  |  |  | 99 |
| LCT Max |  | 6548 | 1631 |  |  |  | 88 |
| NLCT Min |  | 158 | 69 |  |  |  | 56 |
| LCT Min |  | 444 | 419 |  |  |  | 46 |
| NLCT Median |  | 459 | 338 |  |  |  | 73 |
| LCT Median |  | 952 | 736 |  |  |  | 65 |
| Mann-Whitney U Test p-value |  | 0.2041 | 0.1471 |  |  |  | 0.12114 |
| Fisher's Exact Test | NA |  |  | 0.6776 | 1 | 0.8328 |  |
| *Notes | *Alive/<br>Dead |  |  | *Male/<br>Female | *Caucasian/<br>African-American | *Stage 1 or<br>2/Stage<br>3/Stage 4 |  |
| ** Denotes significance, $p < 0.05$ | | | | | | | |

Abbreviations: LCT, Large Cell Transformation; NLCT, Non-Large Cell Transformation; CTCL, Cutaneous T Cell Lymphoma; SS, Sezary Syndrome; Dx, diagnosis; Max, Maximum; Min, Minimum.

### Pathological Features

All patients included in the study were required to have had at least one histopathologically-confirmed biopsy indicating a diagnosis of SS (including flow-cytometric analysis and/or TCR rearrangement). All patients diagnosed with LCT were required to have had at least one histopathologically-confirmed biopsy demonstrating evidence of LCT. Eight patients had biopsies showing transformed MF following or concurrent with a confirmed SS diagnosis, while 20 patients had only a histopathologically-confirmed diagnosis of SS, non-transformed. Although the analysis was largely restricted to the biopsies showing LCT, certain features, such as folliculotropism or epidermotropism, were recorded if they were present on any biopsy specimens (denoted by ‘pathologic features’ in **Supplemental Table 1**).

### Associations of Clinicopathologic Variables with development of LCT

Clinical variables in patients who underwent transformation that showed statistically significant differences between the two groups (transformed and non-transformed) included mean peak LDH value before LCT (934.3 vs. 386.2 U/L, p = 0.0012), and mean maximum TBSA involvement before diagnosis of LCT (76.9% vs. 50.0%, p = 0.0114). Absolute CD8^+^ cell count in LCT vs. non-LCT groups at diagnosis of Sezary Syndrome (101 vs. 668 cells/cmm, p = 0.0455) was significantly different between the groups. When comparing flow cytometry results at the diagnosis of LCT syndrome (for LCT patients) vs. at the most recent blood draw for patients without LCT, there was a significant difference in the mean percentage of CD8^+^ cells (4.573% vs. 14.12%, p = 0.0385). At most recent collection, mean absolute CD8^+^ cell count in the LCT vs. non-LCT group (60 vs. 476 cells/cmm, p = 0.0074) was significantly different between the two groups.

Pathological variables in patients who underwent transformation that showed statistically significant differences between the two groups included presence of Langerhans cell hyperplasia on any biopsy specimen (p = 0.0171), and presence of ulceration on any specimens (p = 0.0034). Results of the significant differences in clinicopathologic variables between the two groups and full summary statistics can be visualized in **Table 1B**.

**Table 1B:**
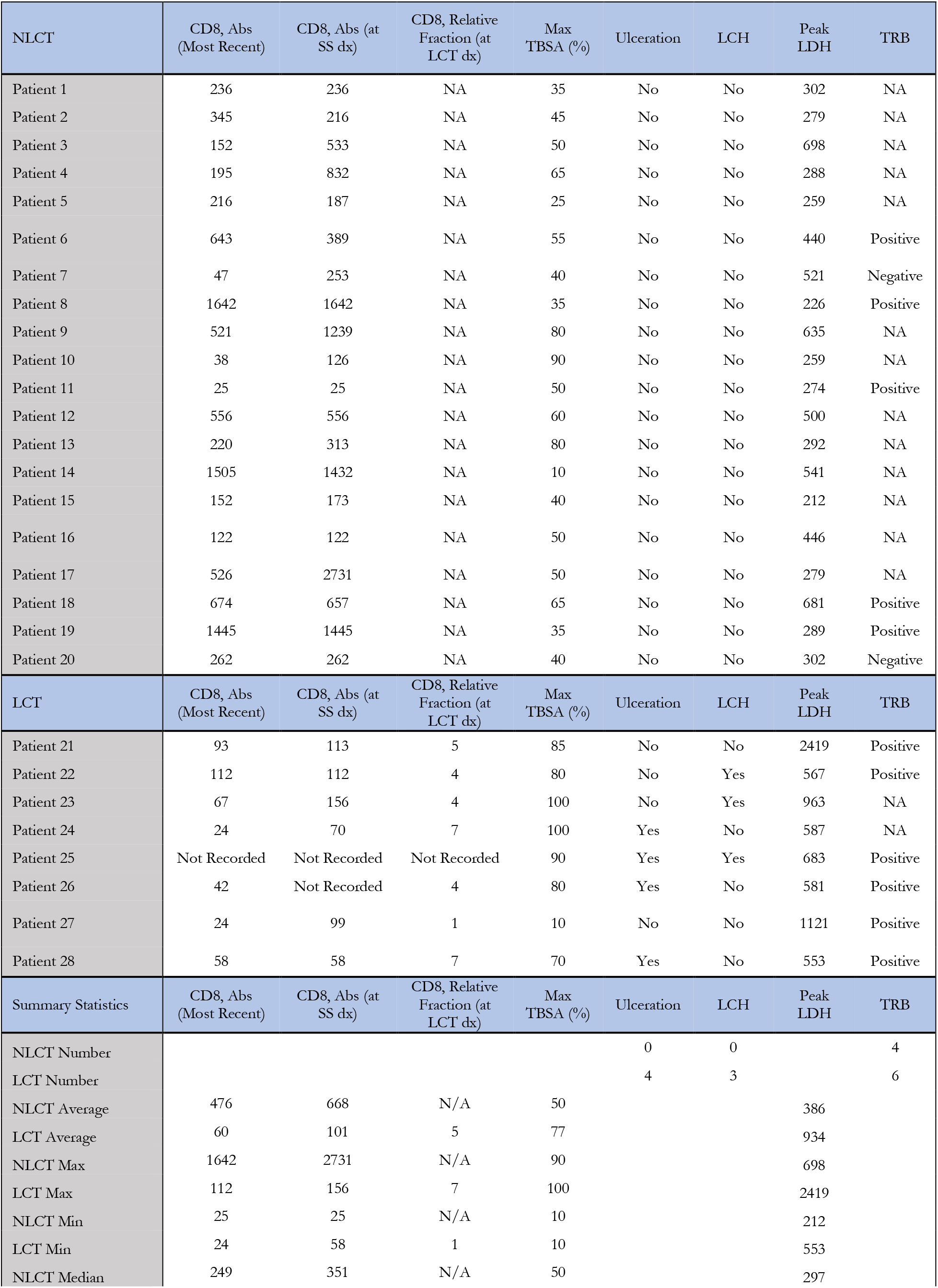
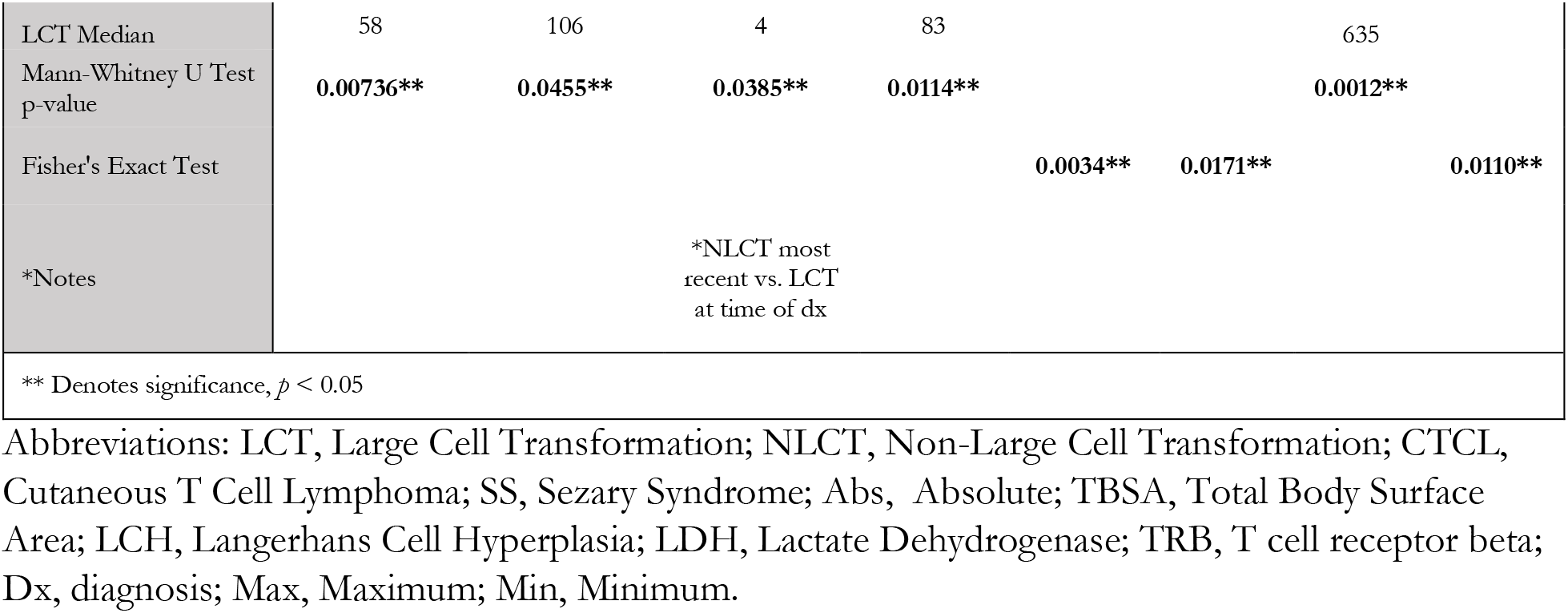
Concise demographic details for each patient included in the study

### Univariate Analysis

Univariate analysis was conducted for each demographic variable and variables found to be significantly different between groups on Mann-Whitney U test and Fisher’s exact test (above). Univariate analyses were conducted for the endpoints of both LCT and OS. Length of SS diagnosis was analyzed as a covariate for both models. Significant variables on univariate analysis for endpoint of LCT (**Figure 2A**) included TBSA (HR 1.043 per unit increase, 95% CI 1.001 – 1.081, p = 0.018), presence of ulceration (HR 10.190, 95% CI 2.339 – 44.393, p = 0.002), presence of Langerhans cell hyperplasia (HR 8.559, 95% CI 1.828 – 40.083, p = 0.006), unit increase in peak LDH before diagnosis of LCT (HR 1.002, 95% CI 1.001 – 1.003, p = 0.002), unit increase in CD8 absolute cell count at diagnosis of Sezary Syndrome (HR 0.988, 95% CI 0.976 – 0.999, p = 0.041), presence of T cell receptor beta clonal rearrangement (HR 16.056, 95% CI 1.902 – 135.568, p = 0.011). On univariate analysis for OS (**Figure 2B**), only Stage of disease was determined to be prognostic for the endpoint, particularly Stage 4 disease vs. Stage 1 or 2 disease (HR 12.811, 95% CI 1.482 – 110.714, p = 0.0205).

**Figure 2:**
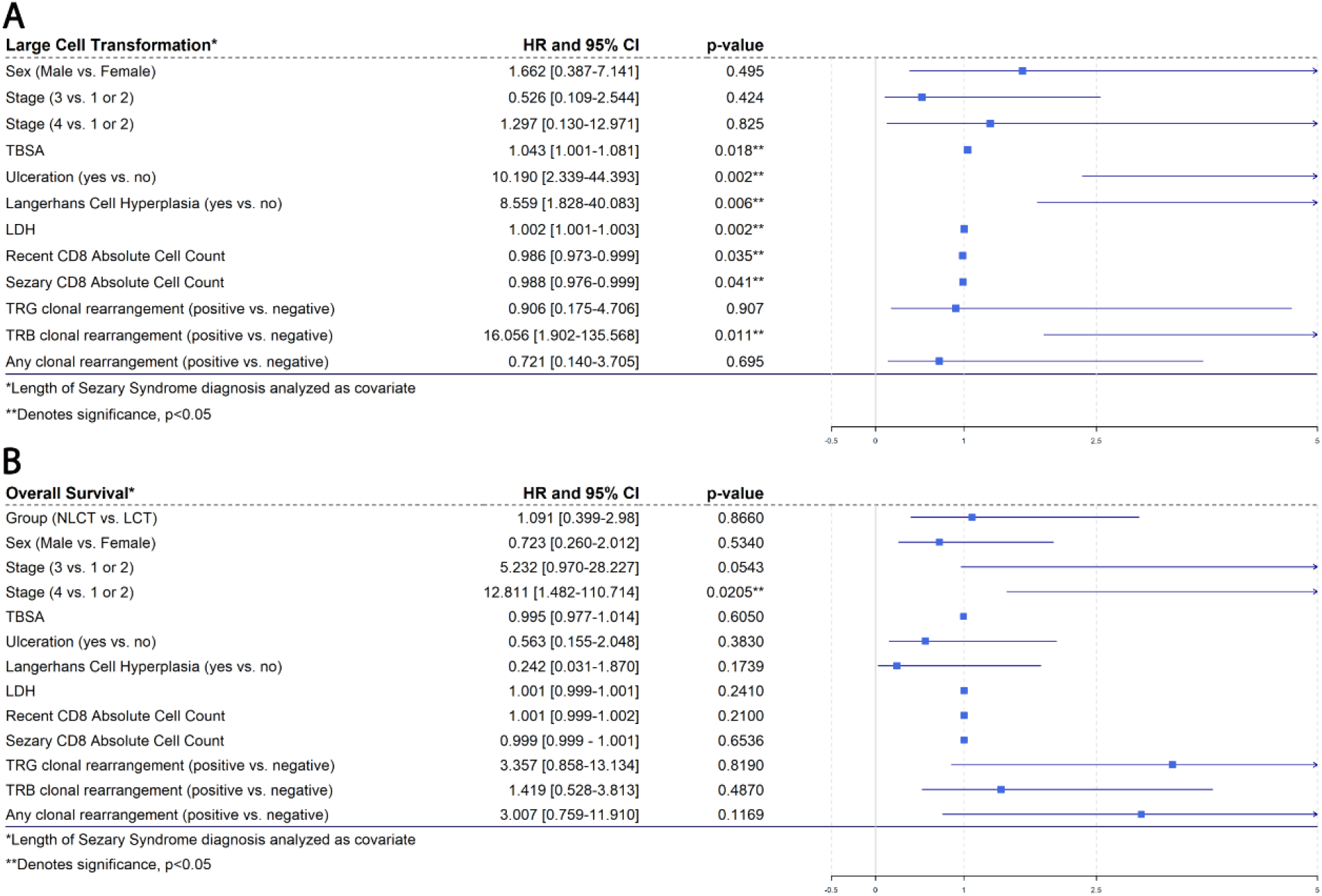
Panel (A) demonstrates univariate analysis for demographic variables and all variables that demonstrated significant differences between the two groups (LCT and NLCT) for the endpoint of LCT. Panel (B) demonstrates univariate analysis for demographic variables and all variables that demonstrated significant differences between the two groups (LCT and NLCT) for the endpoint of OS. Hazard Ratio (HR) and 95% confidence intervals (CI) are noted in the columns with corresponding titles. Significant *p-*values are denoted with two asterisks (**). All univariate analyses were conducted with time from diagnosis of SS as a covariate.

### Survival Analysis

Survival analysis was conducted by taking into account the time to death from both diagnoses of CTCL (**Figure 3A**) and SS (**Figure 3B**) to LCT. There was no significant difference in OS between groups when considering time to diagnosis from MF to LCT (HR 1.679, 95% CI 0.617 – 4.572, p = 0.31) or from SS to LCT (HR 1.367, 95% CI 0.514 – 3.634, p = 0.531). Upon analysis of CD8^+^ T cell counts relative to the median expression of all samples (“High” or “Low”), there were no differences in OS from CTCL diagnosis to death based on relative absolute CD8^+^ T cell count at diagnosis of SS (**Figure 4A**, HR 0.5899, 95% CI 0.2157 – 1.613, p = 0.304) or at most recent blood draw (**Figure 4B**, HR 0.7279, 0.2684 – 1.974, p = 0.533). Additionally, there were no differences in OS from SS diagnosis to death based on relative absolute CD8^+^ T cell count at diagnosis of SS (**Figure 4C**, HR 1.726, 95% CI 0.6275 – 4.748, p = 0.29) or at most recent blood draw (**Figure 4D**, HR 0.9202, 95% CI 0.3359 – 2.521, p = 0.872).

**Figure 3:**
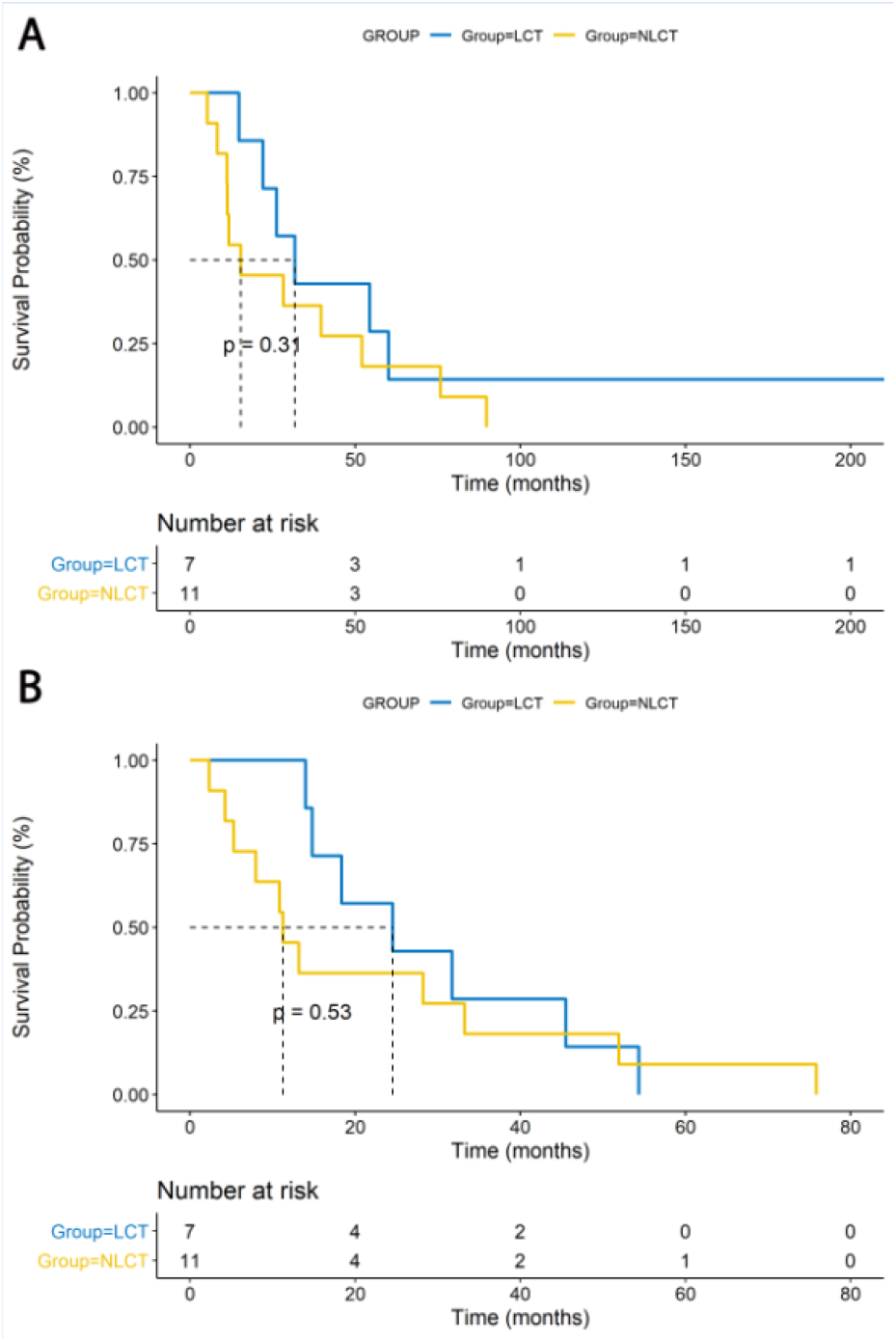
Panel (A) demonstrates the Kaplan-Meier curves displaying differences in time from MF dx to death for patients who underwent large cell transformation (LCT) and patients who did not undergo large cell transformation (NLCT) (HR 1.679, 95% CI 0.617 – 4.572, p = 0.31). Panel (B) demonstrates the Kaplan-Meier curves displaying differences in time from SS dx to death for patients who underwent large cell transformation (LCT) and patients who did not undergo large cell transformation (NLCT) (HR 1.367, 95% CI 0.514 – 3.634, p = 0.531).

**Figure 4:**
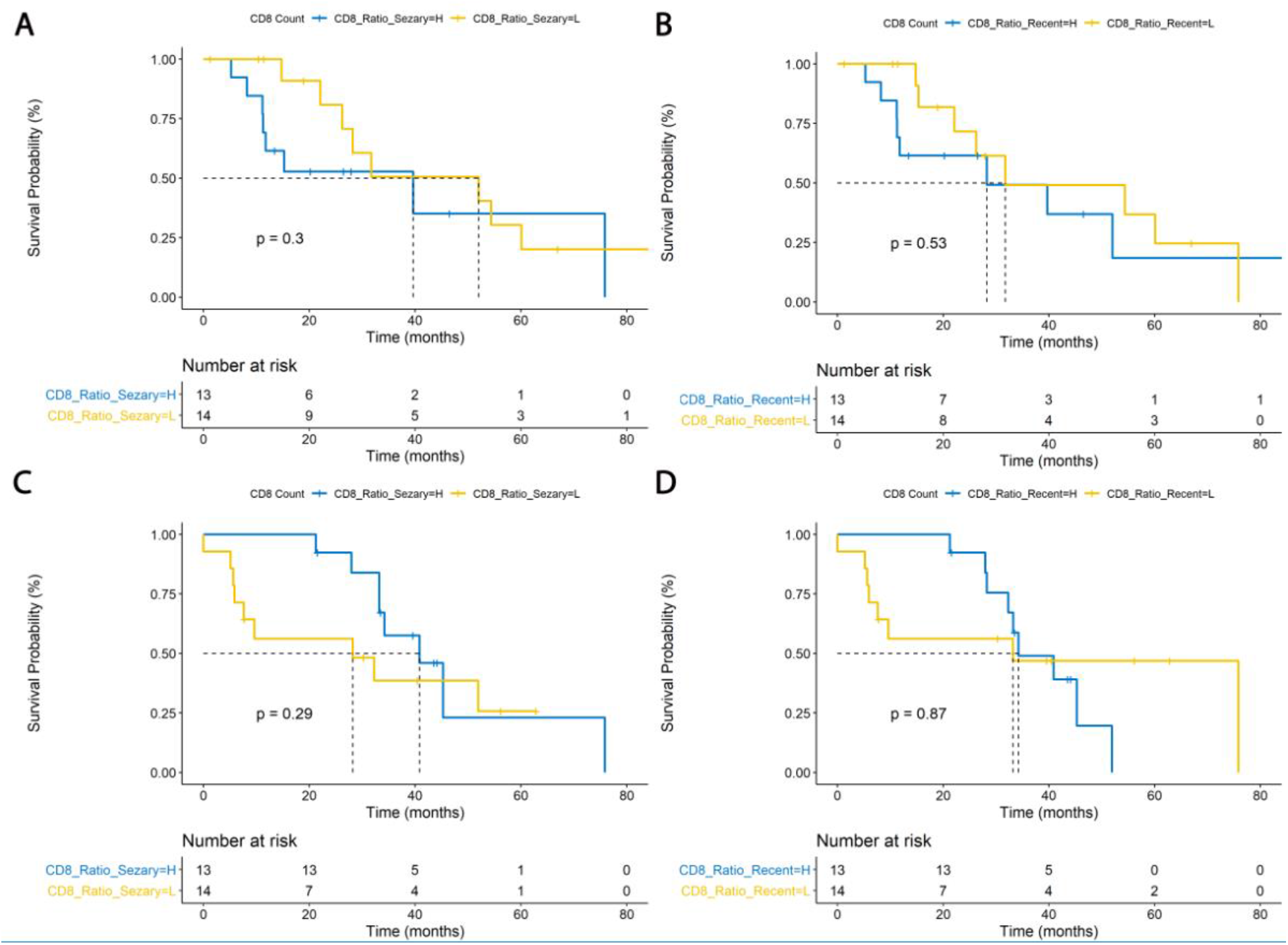
Panel (A) demonstrates a Kaplan-Meier curve for overall survival from initial cutaneous T cell lymphoma diagnosis to death, based on absolute CD8^+^ T cell count at dx of Sezary Syndrome (HR 0.5899, 95% CI 0.2157 – 1.613, p = 0.304). Panel (B) demonstrates a Kaplan-Meier curve for overall survival from initial cutaneous T cell lymphoma diagnosis to death, based on absolute CD8^+^ T cell count at most recent blood draw (HR 0.7279, 0.2684 – 1.974, p = 0.533). Panel (C) demonstrates a Kaplan-Meier curve for overall survival from Sezary Syndrome diagnosis to death, based on absolute CD8^+^ T cell count at dx of Sezary Syndrome (HR 1.726, 95% CI 0.6275 – 4.748, p = 0.29). Panel (D) demonstrates a Kaplan-Meier curve for overall survival from Sezary Syndrome diagnosis to death, based on absolute CD8^+^ T cell count at most recent blood draw (HR 0.9202, 95% CI 0.3359 – 2.521, p = 0.872). All hazard ratios are based on comparison of absolute CD8^+^ T cell Low (L) vs. High (H).

## Discussion

Mycosis fungoides (MF) and Sézary syndrome (SS) represent the most common types of primary cutaneous T-cell lymphoma [2]. Sezary syndrome is a leukemic variant of CTCL [11]. Although MF usually has an indolent clinical behavior, SS is associated with poor outcome [2, 12]. Both can undergo large cell transformation (LCT), a pathologic process wherein neoplastic small lymphocytes transform to a large cell phenotype [5]. Transformation of MF/SS is associated with an increase in clinical aggressiveness [8]. LCT has been confirmed as an independent poor prognostic marker in multiple previous studies [5, 6, 8, 13–18]. “The median time from diagnosis of MF/SS to transformation is usually less than 2 years” [8].

Very few studies have explored the value of specific factors for predicting which patients with MF/SS will eventually undergo LCT [8]. Studies that have researched the clinical outcomes of LCT have determined that advanced stage at the time of transformation diagnosis is associated with poor prognosis [5, 8]. In addition, LCT at initial diagnosis of MF is correlated with decreased survival [5, 6, 8]. Elevated LDH has also been recognized as another independent poor prognostic marker for survival in patients with advanced-stage SS/MF [11] and large cell transformation [5]. Furthermore, Novelli et al. found that a TBSA> 50% involvement by CTCL was significantly associated with a decreased survival [19].

Our study suggests that there are clinical and pathologic factors that may predict the development of LCT. These included greater maximum TBSA with identifiable disease before the diagnosis of LCT, greater maximum LDH level before the diagnosis of LCT, presence of ulceration and Langerhans cell hyperplasia, decreased absolute CD8^+^ cell count at time of SS diagnosis and at the most recent blood draw, and presence of TRB clonal rearrangement.

Increased extent of skin lesions (increased TBSA involvement) and stage of disease at diagnosis have previously been surmised to predict reduced disease-specific survival (DSS) and OS in patients with transformed MF [20]. Our study demonstrates reduced OS for patients with SS and advanced stage at diagnosis as well, while TBSA is predictive of LCT. Both of these parameters are seen as measures of disease aggression. Therefore, TBSA may be a surrogate for disease aggression, rather than a modifiable disease factor, that is correlated with LCT. The same may be said for the presence of ulceration on pathologic specimens. Although T cell receptor beta rearrangement was determined to be prognostic based on analysis of positive results in both groups, the rearrangement was tested for in 7/20 NLCT patients and 6/8 LCT patients, which may be due to a testing bias for patients who were perceived to have more aggressive disease.

Dendritic cells play a pivotal role in linking adaptive and innate immune responses [21]. Although they generally serve to activate effector and helper T cells, leading to downstream anti-tumoral properties, this may not always be the case. In fact, a key mechanism in tumor immune evasion is the modulation of dendritic cell function by tumors and specific tumor-associated factors in the tumor microenvironment. Therefore, despite their abundance in the tumor microenvironment, and their anti-tumoral potential, tumor-infiltrating dendritic cells may often display ineffective and impaired function, and may even mediate immunosuppression [22, 23]. The role of Langerhans cells, a type of dendritic cell, in the pathogenesis and progression of MF/SS is not yet understood [24, 25]. The number of Langerhans cells in MF skin lesions has been reported to be especially increased during the tumor stage than in the patch/plaque stage [24]. Thus, previous studies have suggested that Langerhans cells may enhance tumor progression and trigger malignancy [24–26]. However, other studies have found that Langerhans cell hyperplasia increases survival in MF/SS, and have suggested that Langerhans cells play a defense role against MF/SS [27]. Our study suggests that hyperplasia of Langerhans cells may lead to tumor progression in patients who have been diagnosed with SS, eventually leading to LCT. It is possible that cutaneous T cell lymphomas impair the function of dendritic cells, leading them to act as pro-tumoral, rather than anti-tumoral agents. Further investigation is needed to validate this hypothesis.

It is well-accepted that CD8^+^ T cells play a very important role in mediating anti-tumor immunity, mediated by elimination of tumor cells through recognition of tumor-associated antigens. In cancers such as colorectal, lung, breast, melanoma and glioblastoma, the infiltration of T cells, and especially CD8^+^ T cells, is correlated with better prognosis [28, 29]. There are promising immunotherapies in development and trials for various cancers that aim to boost CD8^+^ T cell-mediated anti-tumor immunity, which include adaptive cell transfer of tumor-reactive T cells (either native or engineered to express tumor specific T cell receptors or chimeric antigen receptors), dendritic cell cancer vaccines (DCVax), and immune checkpoint blockades (including anti-PD-1, anti-PD-L1, and anti-CTLA-4 antibodies) [30]. Our specific analysis poses an interesting question concerning the role of these non-neoplastic cells in cutaneous T cell lymphomas. It has previously been demonstrated that a higher CD8^+^ T cell infiltrate is correlated with better prognosis in patients across plaque-stage MF, tumor stage MF, and CD30-PCTCL, and is correlated with disease aggression [31]. These results are supported also by Hoppe et al. [32] who describe a relationship between higher CD8^+^ T cell proportions and better survival in MF. Our study demonstrated the stark contrast in absolute CD8+ T cell number in the peripheral blood between patients in the LCT and NLCT groups, with significantly decreased absolute counts in patients who underwent transformation, both before and after diagnosis of LCT. This observation may indicate that decreased peripheral CD8+ T cell count may also allow for persistence or continued resistance against LCT. This appears to be in agreement with the demonstrated role of CD8^+^ T cells as anti-tumoral effector cells. Given the retrospective nature of the study, we were unable to obtain sequencing data for the markers of T cell activation that may have offered a more robust analysis of the functional capabilities of the T cells present in the peripheral blood. Further in vitro and in vivo studies are needed to understand these results in a mechanistic context.

Our study is not without limitations. This is a retrospective analysis, and therefore subject to the limitations associated with retrospective study design. Additionally, due to the low prevalence of the disease, our sample size was small, which may explain some of the unusually high hazard ratios seen in our results, and additionally may affect the interpretation of results, such as TRB clonal rearrangement, explained previously. On the other hand, this may mean that larger studies would reveal patterns that we were not able to appreciate with a small number of patients. This limitation may be difficult to overcome, and in vitro and in vivo laboratory experimentation may be required confirm the validity of our hypothesized mechanisms.

In conclusion, our study is the largest study to date that the authors have identified to analyze the risk factors for LCT in patients diagnosed with SS. Here, we identify clinical and pathologic factors that may confer prognostic value for the incidence of LCT, and assess their impact on OS. These include maximum TBSA %, peak LDH, presence of ulceration, TRB clonal rearrangements, Langerhans cell hyperplasia, and decreased levels of CD8^+^ cells in the peripheral blood. Although many of these may be clinical surrogates for the aggressiveness of the patient’s disease, Langerhans cell hyperplasia and absolute CD8^+^ cell counts in the peripheral blood may provide interesting insights and suggest novel therapeutic targets. While Langerhans cells may be playing a pro-tumoral role in patients with SS, greater absolute CD8^+^ cell count in the peripheral blood may serve as a protective factor against the development of LCT. Furthermore, lesser numbers of CD8^+^ T cells may allow for persistence of the disease. Therapy targeted at increasing the number of CD8^+^ T cells in the peripheral circulation may serve to protect patients from developing transformation of their SS. These results indicate the need for further study into the mechanisms of tumor progression based on immune cell populations in patients with SS and CTCL.

## Data Availability

All data is available at request.

## Acknowledgement

TT, YW, LCT are supported by the Michigan Medicine-Peking University Health Science Center Joint Institute for Translational and Clinical Research award. LCT is supported by the NIH (K01-AR072129), the Dermatology Foundation, and the National Psoriasis Foundation.

